# Quantifying contact patterns: development and characteristics of the British Columbia COVID-19 population mixing patterns survey (BC-Mix)

**DOI:** 10.1101/2021.08.10.21261872

**Authors:** Prince A. Adu, Mawuena Binka, Bushra Mahmood, Dahn Jeong, Terri Buller-Taylor, Makuza Jean Damascene, Sarafa Iyaniwura, Notice Ringa, Héctor A. Velásquez García, Stanley Wong, Amanda Yu, Sofia Bartlett, James Wilton, Mike A. Irvine, Michael Otterstatter, Naveed Z. Janjua

**Affiliations:** British Columbia Centre for Disease Control, Vancouver, British Columbia, Canada; School of Population and Public Health, University of British Columbia, Vancouver, British Columbia, Canada; Department of Medicine, University of British Columbia, Vancouver, British Columbia, Canada; Department of Mathematics and Institute of Applied Mathematics, University of British Columbia, Vancouver, British Columbia, Canada; Faculty of Health Sciences, Simon Fraser University, Burnaby, British Columbia, Canada; Department of Pathology and Laboratory Medicine, Vancouver, British Columbia, Canada

## Abstract

**Introduction:** Several non-pharmaceutical interventions such as physical distancing, hand washing, self-isolation, and schools and business closures, were implemented in British Columbia (BC) following the first laboratory-confirmed case of coronavirus disease 2019 (COVID-19) on January 26, 2020, to minimize in-person contacts that could spread infections. The BC COVID-19 Population Mixing Patterns survey (BC-Mix) was established as a surveillance system to measure behaviour and contact patterns in BC over time to inform the timing of the easing/re-imposition of control measures. In this paper, we describe the BC-Mix survey design and the demographic characteristics of respondents.

**Methods:** The ongoing repeated online survey was launched in September 2020. Participants are mainly recruited through social media platforms (including Instagram, Facebook, YouTube, WhatsApp). A follow up survey is sent to participants two to four weeks after completing the baseline survey. Survey responses are weighted to BC’s population by age, sex, geography, and ethnicity to obtain generalizable estimates. Additional indices such as the material and social deprivation index, residential instability, economic dependency, and others are generated using census and location data.

**Results:** As of July 26, 2021, over 61,000 baseline survey responses were received of which 41,375 were eligible for analysis. Of the eligible participants, about 60% consented to follow up and about 27% provided their personal health numbers for linkage with healthcare databases. Approximately 50% of respondents were female, 39% were 55 years or older, 65% identified as white and 50% had at least a university degree.

**Conclusion:** The pandemic response is best informed by surveillance systems capable of timely assessment of behaviour patterns. BC-Mix survey respondents represent a large cohort of British Columbians providing near real-time information on behavioural and contact patterns in BC. Data from the BC-Mix survey would inform provincial COVID-19-related control measures.

## Introduction

The novel coronavirus disease 2019 (COVID-19), caused by the severe acute respiratory syndrome coronavirus 2 (SARS-CoV-2), has spread worldwide since December 2019. A global pandemic was declared by the World Health Organization in March 2020 and, as of July 2021, there have been over 200 million cases of COVID-19 infections and over 4.3 million resultant deaths globally (1). As vaccine rollouts continue at varying rates worldwide, physical distancing measures (2) remain among the most effective methods for COVID-19 prevention and control (3). Many governments have put in place physical distancing measures such as travel restrictions, closure of schools and workplaces, and the banning of large group gatherings to interrupt the transmission of SARS-CoV-2. These measures attempt to reduce contact between infected and healthy individuals in order to minimize disease spread and the impact on the healthcare system.

British Columbia (BC) is located on the West Coast of Canada and covers almost a million square kilometres. It has a diverse population of approximately 5.15 million as of July 1, 2020 (4). Public health officials in BC began urging the public to practice physical distancing and avoid any non-essential travel in early March 2020. By March 17, 2020, a public health emergency was declared in the province and various physical distancing measures were implemented (5). These included restriction of indoor and outdoor gatherings, closure of businesses that were unable to meet physical distancing measures, self-isolation requirements after travelling outside the country, and general physical distancing in all public space. While these measures were important for controlling the rapid spread of disease, they also had sweeping economic, social, and mental health impacts.

Assessing the impact of physical distancing measures on person-to-person contact can provide valuable information for refining control measures and help minimize both COVID-19-related disease burden and the related economic, social, and mental health impacts. Early detection of COVID-19 resurgences requires mechanisms for tracking precursors of transmission, including changes in social contacts, mixing patterns and physical distancing behaviours as well as early signals of a COVID-19 spread. Although methods such as mathematical modelling can estimate the potential for resurgences, these methods often lack population-based empirical data on contact patterns, especially on the varying levels of contact patterns exhibited by different demographic groups in the population. These population-specific data could better inform mathematical models by incorporating explicit knowledge of contact patterns that are driving transmission rather than inferring these from reported cases and hospitalizations (6,6). Ultimately, they serve as an evidence-base to guide targeted measures that are amenable to actions by the government to ensure that the COVID-19 cases remain below the resurgence thresholds.

Various studies have assessed the impact of physical distancing measures imposed by governments on local contact patterns and behaviours during the COVID-19 pandemic in Belgium (7), Greece (8), Kenya (9), Luxembourg (10), the Netherlands (11), and the U.K (12). Such surveys can measure the public’s compliance with the physical distancing measures and provide valuable information to inform other public health measures that may be necessary to avoid further waves of COVID-19 infections. In addition, the impact of physical distancing measures on mixing patterns and contact behaviours may vary across different age groups, and by individuals’ primary place of activity such as schools or workplaces (8,13–15).

Here, we describe the development the BC COVID-19 Population Mixing Patterns survey (BC-Mix), an ongoing online survey to monitor and assess social contact behaviours and mixing patterns in BC, Canada, during the COVID-19 pandemic. We detail the development of the survey and recruitment of respondents, as well as the characteristics of the participants.

## Methods and analysis

### Survey design and methodology

The BC-Mix (http://www.bccdc.ca/our-research/projects/bc-mix-covid-19-survey) uses a cross-sectional survey design with longitudinal follow-up. Eligible population include residents of BC who are at least 18 years of age. The survey began on September 4, 2021, and as of August 2021 is ongoing. Once a participant has completed the survey for the first time, they are invited for repeated follow-up. The first-time responses are referred to as the ‘baseline’. Participants responding to the baseline survey are invited to complete the first follow-up survey after two weeks. Subsequent follow-up surveys are then sent in four-week intervals, following the completion of the previous survey.

### Participant recruitment

To capture participants from a broad demographic range, the survey invitation and survey are disseminated through Instagram, Facebook, YouTube, WhatsApp, Twitter, and Google search engine results pages. The Google Ads Audience manager and Facebook Ads manager allow for paid advertisements to be targeted at specific audiences. We use these tools to target the survey advertisement campaigns to only residents of BC who are 18 years and above. We also monitor the demographic profile of survey participants and occasionally use these functions to target recruitment to age groups or sex that may be under-represented (16).

To help capture underrepresented groups, we promote the survey to various ethnic populations. For instance, a South Asian community organization promotes the survey on their social media pages and also sends the survey to individuals on their mailing list. Although the survey is in English, it is also promoted in different languages (specifically, Korean and Farsi) to members of minority community groups in BC on their social media pages. Flyers are also distributed at grocery stores and restaurants particularly including those frequented by minority groups.

### Participant and public involvement

The initial version of the BC-Mix survey was first piloted with a randomly selected sample of the BC population and feedback received was incorporated in the final version before the official launch. Methods of recruitment and priority of research questions were also informed by discussions with members of the public and with a community group. We also receive input from survey participants on an ongoing basis through a dedicated e-mail address. We plan to create dashboards and other infographics of the study results on the study website. A newsletter suitable for non-specialist audience will also be sent to participants.

### Survey domain and case definitions

The BC-Mix survey instrument was adapted from the POLYMOD study (14) and the Berkeley Interpersonal Contact Study [BICS] (17) and was administered through Qualtrics (18), an online survey tool. The baseline survey comprises 94 questions across six key domains:

1. **Demographic information:** This domain includes age, sex, gender, ethnicity, education, employment, household characteristics, and postal code.
2. **COVID-19 testing and results, symptoms, and health behaviours:** This domain captures COVID-19 testing information, symptoms, and behaviours such as doctor visits following symptoms.
3. **Activities and behaviour in and outside of the home:** This domain captures social contact and mixing behaviours such as number of contacts, location, and duration of contact during the past 24 hours. Other questions in this domain include age and sex of contact, and relationship of respondent to the contact persons, physical distancing behaviour (e.g., handwashing) and personal protective equipment use. Initially, respondents were asked to provide this information for up to three of their reported contacts. We began collecting data for up to 10 contacts from December 11, 2020. Also from December 11, 2020, we began collecting general information about greater than 10 contacts i.e., if a participant reports more than 10 contacts per day, they are asked general questions about these contacts for e.g., age group, duration, and location of the majority of those contacts. If majority of contacts took place at a workplace setting, a follow up question asks respondents to report the type of work setting where the contacts occurred.
4. **Internet and social media use:** This domain captures information on internet and social media use in terms of most frequently used platform and frequency of use.
5. **Perceptions and attitudes around COVID-19:** This domain measures the respondent’s perception of the physical distancing measures, and their self-confidence or ability to carry out them.
6. **COVID-19 vaccine acceptance sub-questionnaire:** This sub-questionnaire was added on March 8, 2021. Items from this domain were developed using a vaccine acceptance behavioral framework, which synthesizes constructs from the Theory of Reasoned Action (TRA)(19), Theory of Planned Behavior (TPB)(20,21) and the Health Belief Model (HBM) (22), to understand and predict the uptake of COVID-19 vaccine. According to the TRA, the best single predictor of behaviour is an individual’s intention (23). Intentions, in turn, are an outcome of the individual’s attitude toward performing the behavior in question, and/or the individual’s perceptions of support from family and friends (subjective norms) for engaging in the behavior (24). Perceived control or self-efficacy, the confidence that one has the ability to perform the intended behavior (25), is another important construct taken from TPB. The TPB assumes that an individual’s perception of whether they can successfully engage in a particular behavior often has a direct effect on their intentions, such as getting a vaccine (26). The widely-used HBM, has previously been used to evaluate beliefs and attitudes toward seasonal influenza and pandemic swine flu vaccines as well as the COVID-19 vaccine (27–29). Relevant constructs from HBM were applied to develop questionnaire items to assess perceived threat of contracting the COVID-19, perceived severity of disease if infected and belief in the safety and effectiveness of getting the vaccine. Overall, this sub-questionnaire is meant to provide an understanding of some of the individual level health beliefs, perceptions and attitudes that may influence vaccine uptake. The vaccine acceptance sub-questionnaire has the following the domains: *Attitude (perceived susceptibility, severity, benefits and barriers), Descriptive and Subjective Norms, Perceived Control* and *Intention*.

Location data is used to generate other indicators at the area level. For example, the Quebec Material and Social Deprivation combines six indicators related to health and welfare that represent material or social deprivation based on Canadian Census data, including 1) proportion of persons without high school diploma 2) ratio of employment to population 3) average income 4) proportion of persons separated, divorced, widowed 5) proportion of single-parent families and 6) proportion of people living alone (30).

A full list of key variables in the survey and definitions is presented in S1 Table in the Supplementary file.

### Analysis, data cleaning and weighting

A survey completion rate of at least 33% of questions, valid non-missing responses for the sex and age questions are required for inclusion for weighting the survey data and further analysis. All duplicates are removed.

To ensure that the BC-Mix sample is representative of the BC population, survey data are weighted to obtain generalizable estimates (Table 1). Using the 2016 Census data (31), the survey is weighted with the following auxiliary variables: age, sex, geography, and ethnicity using the weighting adjustment technique (32) in the following hierarchy: As our first criterion, we consider age, sex, geography and ethnicity as our auxiliary variables. If a record has valid responses for all these variables except the ethnicity variable, then the survey weight is generated using only age, sex, and geography (second criterion). If a record does not meet the first and second criteria, then we apply the third criterion which uses age, sex, and ethnicity as the auxiliary variables. Finally, we use only age and sex as auxiliary variables if a record does not satisfy the first three criteria.

**Table 1.** Participant profile of BC-Mix baseline data (n=41,375), September 04, 2020-July 26, 2021

|  |  | Survey |  |  |  | British Columbia population |  |
| --- | --- | --- | --- | --- | --- | --- | --- |
|  |  | Un-weighted frequency | Un-weighted % (excl. missing) | Weighted frequency | Weighted % | Population frequency | Population % |
| Sex | Male | 6,823 | 16.5 | 21,293 | 50.0 | 1,805,105 | 48.5 |
|  | Female | 34,552 | 83.5 | 21,261 | 50.0 | 1,914,755 | 51.5 |
|  | Missing |  |  |  |  |  |  |
| Age | 18-34 | 4,978 | 12.0 | 11,575 | 27.2 | 1,002,745 | 27 |
|  | 35-54 | 12,110 | 29.3 | 14,194 | 33.4 | 1,251,835 | 33.7 |
|  | 55+ | 24,287 | 58.7 | 16,784 | 39.4 | 1,465,280 | 39.4 |
| Race/ethnicity | Indigenous | 1,757 | 4.4 | 2,180 | 5.3 | 186,705 | 5 |
|  | Chinese | 882 | 2.2 | 4,451 | 10.9 | 418,035 | 11.2 |
|  | White | 35,026 | 87.5 | 26,383 | 64.6 | 2,448,155 | 65.8 |
|  | South Asian | 606 | 1.5 | 3,473 | 8.5 | 280,470 | 7.5 |
|  | Other | 1,766 | 4.4 | 4,352 | 10.7 | 386,495 | 10.4 |
|  | Missing/Unknown | 1,338 | n/a | 1715 | n/a | n/a | n/a |
| Health region | Fraser Health | 8,451 | 26.1 | 11,793 | 36.2 | 1,347,410 | 36.2 |
|  | Interior Health | 6,143 | 19.0 | 5,336 | 16.4 | 595,105 | 16 |
|  | Northern Island | 1,825 | 5.6 | 1,828 | 5.6 | 213,235 | 5.7 |
|  | Vancouver Coastal | 7,315 | 22.6 | 8,118 | 24.9 | 934,055 | 25.1 |
|  | Vancouver Island | 8,640 | 26.7 | 5,535 | 17.0 | 630,055 | 16.9 |
|  | Missing/Unknown | 9,001 | n/a | 9,943 | n/a | n/a | n/a |
| Education | Below high school | 807 | 2.5 | 1,096 | 3.0 | 2,301,030 | 12.5 |
|  | Below bachelor | 16,928 | 51.7 | 15,176 | 47.0 | 466,295 | 61.9 |
|  | University degree | 15,029 | 45.9 | 16,273 | 50.0 | 952,535 | 25.6 |
|  | Missing/Unknown | 8,611 | n/a | 10,009 | n/a | n/a | n/a |
| Employment status | Employed full-time (30 hours or more/week) | 10,654 | 32.0 | 13,608 | 40.8 | n/a | n/a |
|  | Employed part-time | 2,993 | 9.0 | 3,131 | 9.4 | n/a | n/a |

| Survey |  |  |  |  | British Columbia population |  |
| --- | --- | --- | --- | --- | --- | --- |
|  | Un-weighted frequency | Un-weighted % (excl. missing) | Weighted frequency | Weighted % | Population frequency | Population % |
| Self-employed | 2,704 | 8.1 | 3,013 | 9.0 | n/a | n/a |
| Unemployed but looking for a job | 952 | 2.9 | 1,522 | 4.6 | n/a | n/a |
| Unemployed and not looking for a job | 406 | 1.2 | 510 | 1.5 | n/a | n/a |
| Full-time parent, homemaker | 879 | 2.6 | 740 | 2.2 | n/a | n/a |
| Retired | 12,757 | 38.3 | 8,096 | 24.3 | n/a | n/a |
| Student/Pupil | 566 | 1.7 | 1,197 | 3.6 | n/a | n/a |
| Long-term sick or disabled | 968 | 2.9 | 914 | 2.7 | n/a | n/a |
| Prefer not to answer | 424 | 1.3 | 619 | 1.9 | n/a | n/a |
| Missing/Unknown | 8,072 | n/a | n/a | n/a | n/a | n/a |
| Material Deprivation Index |  |  |  |  |  |  |
| 1 (Privileged) | 6,407 | 22.3 | 6,100 | 21.8 | n/a | n/a |
| 2 | 6,475 | 22.5 | 5,873 | 21.1 | n/a | n/a |
| 3 | 6,972 | 24.2 | 6,010 | 21.6 | n/a | n/a |
| 4 | 4,822 | 16.8 | 5,187 | 18.7 | n/a | n/a |
| 5 (Deprived) | 4,085 | 14.2 | 4,656 | 16.8 | n/a | n/a |
| Missing | 1,2614 | n/a | n/a | n/a | n/a | n/a |
| Social Deprivation Index |  |  |  |  |  |  |
| 1 (Privileged) | 4,932 | 17.2 | 5,199 | 17.9 | n/a | n/a |
| 2 | 4,756 | 16.5 | 4,752 | 16.4 | n/a | n/a |
| 3 | 6,311 | 21.9 | 6,022 | 20.8 | n/a | n/a |
| 4 | 5,932 | 20.6 | 6,054 | 20.9 | n/a | n/a |
| 5 (Deprived) | 6,830 | 23.8 | 6,957 | 24.0 | n/a | n/a |
| Missing | 1,2614 | n/a | n/a | n/a | n/a | n/a |
| Follow up consent |  |  |  |  |  |  |
| Yes | 20,633 | 63.8 | 19,051 | 58.9 | n/a | n/a |
| No | 11,689 | 36.2 | 13,275 | 41.1 | n/a | n/a |
| Missing | 9,053 | n/a | n/a | n/a | n/a | n/a |
| Data linkage consent |  |  |  |  |  |  |
| Yes | 7,290 | 27.3 | 7,318 | 26.4 | n/a | n/a |

|  | Survey |  |  |  | British Columbia<br>population |  |
| --- | --- | --- | --- | --- | --- | --- |
|  | Un-weighted<br>frequency | Un-weighted %<br>(excl. missing) | Weighted<br>frequency | Weighted<br>% | Population<br>frequency | Populatio<br>% |
| No | 19,467 | 72.8 | 20,362 | 73.6 | n/a | n/a |
| Missing | 14,618 | n/a | n/a | n/a | n/a | n/a |

Survey weights are estimated separately for baseline and for each follow-up. To assess participant profile, we computed un-weighted and weighted frequency and percentages of key demographic variables using SAS Software version 9.4. Baseline survey data was used to provide the survey participant profile and in comparison, with BC population profile (Table 1). To assess potential systematic differences between eligible and ineligible responses, a comparison of the baseline eligible participants versus ineligible participants is presented the S2 Table in the Supplementary file. Participant profile of follow up surveys is also presented in S3 Table in the Supplementary file.

### Ethics

Informed consent was sought on the survey start page. The study was reviewed and approved by the University of British Columbia Behavioral Research Ethics Board (No: H20-01785).

## Results

As of July 26, 2021, there were 61,183 respondents who participated in the baseline survey of which 41,375 were eligible for analysis. There were 15,194 (eligible=10,993) participants in the first follow-up survey, 11,343 (eligible n=8,164) in the second, 8,521 (eligible n=6,375) in the third, 6,487 (eligible n=4,981) in the fourth, 5,014(eligible=3,891) in the fifth, 4,094 (eligible=3,184) in the sixth, 3,125 (eligible n= 2,417) in the seventh and 2,317 (eligible n=1,760) participants in the eighth follow-up survey (Fig. 1).

**Fig. 1.**
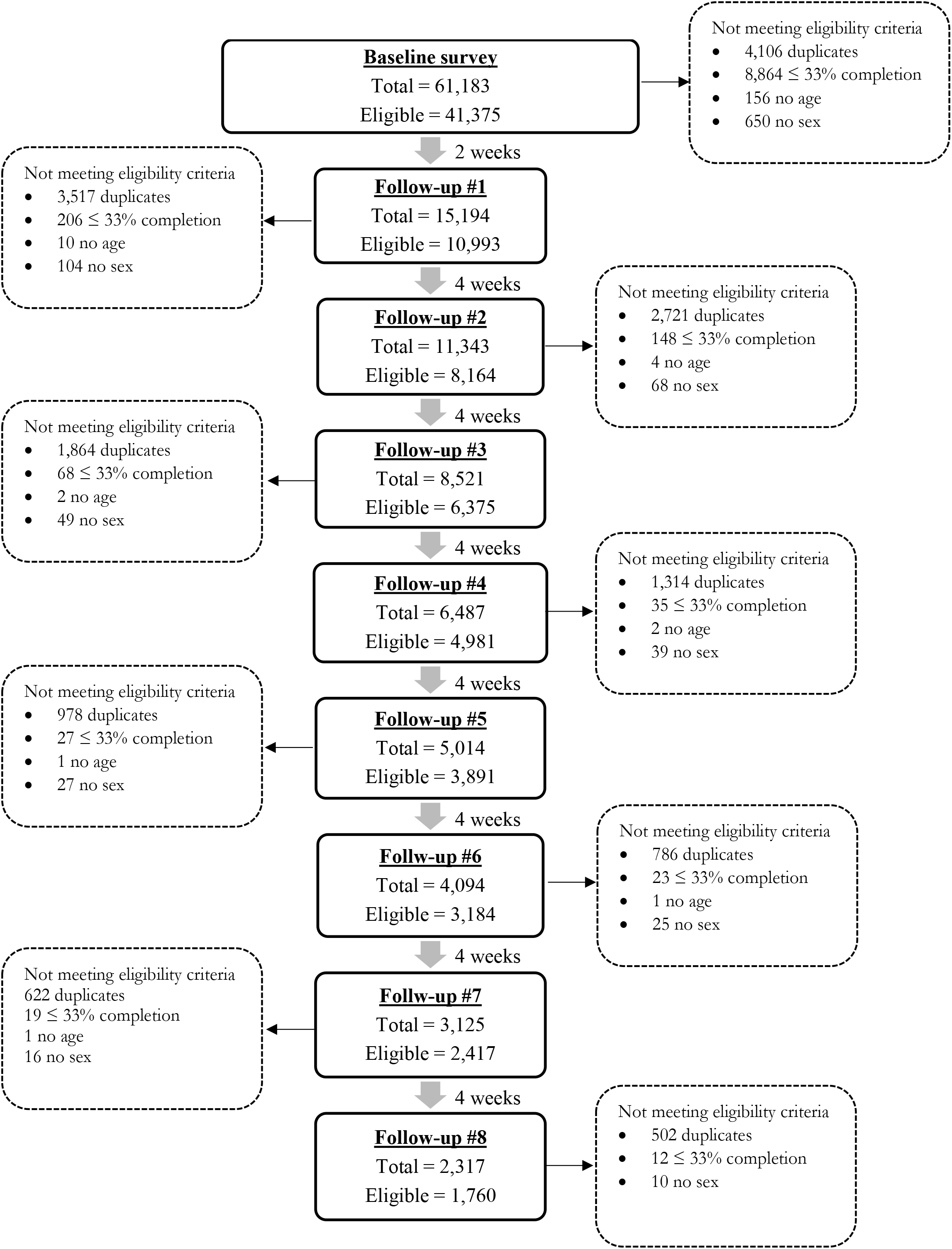
Participant flowchart

Considering the baseline sample (Table 1), there were approximately equal number of male and female (weighted % of female =50.0%). Majority of participants were 55 years or older (weighted %= 39.4%), self identified as White (weighted %= 64.6%), had at least a university degree (weighted %= 50.0%) and lived in the Fraser Health region (weighted %= 36.2%).

Almost 63.8% (unweighted n=20,633) consented to a follow-up after the baseline survey and at least 94.2% (unweighted n=10,357) consented to receiving subsequent follow-up surveys (Table 1 and S3 Table in the Supplementary File). Approximately 27.3% (unweighted n=7,290) of respondents in the baseline provided their personal health numbers for linkage with other healthcare utilization databases.

After weighting, the baseline survey sample is representative of the general BC population in terms of age, sex, health region, and ethnicity (Table 1). The distribution of the eligible participants was also similar to the distribution of ineligible participants in terms of sex, age, race/ethnicity and geography/health region (S2 Table in the Supplementary file).

## Discussion

Following the identification of COVID-19 cases in BC, several interventions including physical distancing measures were implemented to limit the spread of COVID-19 in the province. Subsequently, the BC-Mix was developed by the BC Centre for Disease Control (BCCDC)(33) as part of an early warning system for monitoring social and physical interactions between individuals of different age-groups and demography, and to help predict when COVID-19 transmission might further increase. This paper describes BC-Mix survey methods and the profile of survey respondents.

Recent studies similar to the BC-Mix have assessed social contact patterns relevant to the spread and control of COVID-19 in different countries(7–12,34,35) many of which have adapted features of the POLYMOD project (14). The 2020 Belgian CoMix survey (7) is an online longitudinal survey that closely monitors changes in social mixing behaviours among a sample of Belgian adults (aged 18 years and above). The U.K CoMix survey assesses contact patterns of a representative sample of U.K adults. Launched on March 24, 2020, participants are followed up every 2 weeks to monitor changes in their self reported behaviours (12). In Canada, the Quebec-based CONNECT study uses population-based survey to assess social contacts and mixing patterns (34). Brankston and colleagues (35) also used paid panel representative of Canadian adults to construct contact patterns to determine the impact of physical distancing measures on COVID-19 transmission. Most of these studies commissioned market research companies or used survey panels to recruit participants (7,12,17,35). While market companies or survey panels offer a convenient approach to sampling, they have some challenges. Panels are made of membership in loyalty programs or other panels constituting a select group of population and may therefore not represent complete random recruitment from a population of interest.

The use of targeted social media advertisement for participant recruitment has gained prominence in health research (16,36), having been applied in areas such as mental health (37), cannabis use (38), smoking behaviour (39) and in other health related studies (40). For our survey, we use social media advertisement and other recruitment strategies. Although social media-based recruitment does not necessarily generate a random sample of the general population given the characteristics of people who are on social media may differ from those who are not, social media channels like Facebook, Instagram, Twitter, and others have powerful targeting capabilities that allow researchers to target advertisements to users with specific demographic characteristics. They also have the advantage of reaching hard-to-reach populations (37–39).

Quota sampling has been used by other studies to achieve representativeness (7,44). We used two approaches to achieve the same goal: adaptive recruitment through promotion and targeting to specific populations and then post hoc weighting. Our survey tool does not set quotas on recruitment but uses targeted advertisements to improve representativeness.

The following issues should be considered for interpretation of results from BC-Mix. Some population groups are underrepresented in the survey possibly due to lack of access to social media. These are people who are economically marginalised and less likely to have access to a computer/electronic device or to have access to the internet/cellular data, e.g., people living in poverty, people who are unemployed, people who are unhoused, etc. Also, people who are in prison (sentenced or on remand) or people who are under immigration detention may not have access to the internet or cellular devices. Our survey responses may be subject to recall bias since we ask respondents to recall contacts and other behaviours or activities from the previous day. Other studies have used diaries (14) to overcome this weakness but this may be logistically challenging and attrition with this method may be quite high. Another potential bias inherent in our survey is the issue of reporting bias, as respondents may respond in ways consistent with the laws around physical distancing. In addition, the BC-Mix is available only in English, thus excluding individuals who cannot communicate in English. This notwithstanding, according to the 2016 Census, 96.6% of BC’s population indicated that they can converse in English (31). Therefore, we do not believe that any bias associated with language would be significant. Another limitation to mention is the large number of recruits that were ineligible and the attrition between successive rounds of survey. This could be related to survey fatigue, or the time required to complete the survey.

Our survey has several strengths. Web-based surveys like the BC-Mix provide timely information for pandemic response (45). Also, during an infectious disease pandemic, web-based surveys offer a more convenient approach to data collection compared to in-person or other modes of data collection. We also found paid advertisements to be more cost effective compared to the cost of panel data from survey companies (36). An additional strength of our study is its large sample size. Our total recruited sample of over 61,000 participants compares to the 1,356 participants in the U.K CoMix study (12), the 9,743 participants in the BICS study (17) study, 1,542 participants in the Belgian CoMix study (7) and the 7,290 participants in the POLYMOD study (14). In addition, because we opted to achieve representativeness post-data collection (at the analysis stage), we were able to consider many important variables besides age and sex in our weighting strategy. It would have been logistically challenging to consider all these variables had we used quota-sampling given that many market research company panels were limited in terms recruitment by age, sex, and geography. Using many auxiliary variables in our weighting strategy increased the representativeness of the BC population.

## Conclusion

To our knowledge, the BC-Mix is the first and largest surveillance tool providing real time quantitative data on mixing patterns and contact characteristics in BC and one of the largest in North America. Tools such as the BC-Mix are integral to the COVID-19 pandemic response to provide critical data to inform the timing of loosening or re-imposition of physical distancing measures. Further analyses on contact patterns, relationship of contact patterns with transmission, disparities in contact patterns, facemask use, are in progress and will be published soon.

## Data Availability

Data available upon request

## Supporting information

S1 Table. BC-Mix variable names and definitions

S2 Table. Comparison of baseline eligible and ineligible participants, frequencies and proportions

S3 Table. Participant profile of BC-Mix follow up surveys: frequencies and proportions (%)

## Authors’ contributions

**Conceptualization:** Naveed Z. Janjua, Prince A. Adu

**Survey design:** Naveed Z. Janjua, Prince A. Adu, Terri Buller-Taylor, Bushra Mahmood

**Data curation:** Prince A. Adu, Amanda Yu, Stanley Wong

**Statistical analysis:** Prince A. Adu

**Funding acquisition:** Naveed Janjua

**Methodology:** Naveed Z. Janjua, Prince A. Adu, Mawuena Binka, Terri Buller-Taylor, Bushra Mahmood, Sarafa Iyaniwura, Michael Otterstatter

**Writing first draft:** Prince A. Adu, Dahn Jeong, Mawuena Binka, Terri Buller-Taylor, Sarafa Iyaniwura, Notice Ringa

**Writing-review & editing:** Naveed Z. Janjua, Héctor A. Velásquez García, Bushra Mahmood, Makuza Jean Damascene, Mawuena Binka, James Wilton, Sofia Bartlett, Michael Otterstatter, Mike Irvine.

## Acknowledgement

The authors will like to express their gratitude to Mei Wong and Dr Joan Hu for their methodological guidance.

## Word Count

3,200 words

