## Supplementary file for "Quantifying contact patterns: development and characteristics of the British Columbia COVID-19 population mixing patterns survey (BC-Mix)"

**S1 Table:** BC-Mix variable names and definitions

| **Domain** | **Variable** | **Definition** |
| --- | --- | --- |
| Demographic information | | |
|  | Age, sex, gender, ethnicity, first name, last name, personal health number, postal code, employment status, education, occupation | Age, sex, gender, ethnicity, first name, last name, personal health number, postal code, employment status, education, occupation of respondent |
|  | Number of adults living in respondent's household | How many adults live in your household? |
|  | Number of children living in respondent's household | How many children (under 18 years) live in your household? |
| Perceptions and attitudes around COVID-19 | | |
|  | Satisfaction with provincial COVID-19 response | How satisfied are you with how COVID-19 has been managed in the province? |
|  | Knowledge of COVID-19 | How much do you know about COVID-19? |
|  | Attitude to COVID-19 #1 | To what extent do you agree or disagree with the following statements? - 1) COVID-19 would be a serious illness for me if I caught it 2) I think I am likely to catch COVID-19 3) If I don’t follow government advice, I might spread COVID-19 to someone who is vulnerable. 4) My boss expects me to work when I am feeling unwell or sick 5) If I could not work because of COVID-19, I would still get paid 6) If I had to isolate myself for 14 days because of COVID-19, I would have enough food and supplies for 14 days 7) If I had to isolate myself for 14 days because of COVID-19... - Someone else would be able to look after my children |
|  | Attitude to COVID-19 #2 | How effective do you think the following are at slowing the spread of COVID-19? -1) Meeting up with fewer people than normal 2) Avoiding crowded spaces 3) Staying at home for 14 days if you have ... - Severe symptoms (e.g., severe cough or high temperature). |
|  | Attitude to COVID-19 #3 | How much, if at all, have you changed the number of face-to-face interactions with other people as a result of the COVID19 pandemic? |
|  | Attitude to COVID-19 #4 | How well do you think you are doing at keeping physically distanced from people outside your home? |
|  | Attitude to COVID-19 #5 | How concerned are you personally about the spread of COVID19? |
| Attitude toward COVID-19 vaccine | | |
|  | Vaccination status (1st or 2nd shot) | Have you received the COVID-19 vaccine (either 1st or 2nd shot)? |
|  | Date of 1st shot | When did you receive your 1st COVID-19 vaccine shot? |
|  | Vaccination status (2nd shot) | Have you received your 2nd COVID-19 vaccine shot? |
|  | Date of 2nd shot | When did you receive your 2nd COVID-19 vaccine shot? |
|  | Perceived risk | I believe I am at risk of becoming infected with COVID-19. |
|  | Perceived susceptibility | With the way my life is, I believe I am at a high risk of getting COVID-19 (e.g., risks at my work, recreational activities, people I live with, etc.) |
|  | Perceived protection 1 | I believe a COVID-19 Vaccine will protect me from getting the virus. |
|  | Perceived protection 2 | I believe a COVID-19 vaccine will decrease my chance of getting seriously ill from COVID-19. |
|  | Trust | I do not trust the COVID-19 vaccine. |
|  | Effectiveness | I am concerned about the effectiveness of the COVID-19 vaccination. |
|  | Safety | I am concerned about the safety of the COVID-19 vaccination. |
|  | Subjective norm 1 | Most of the people I know are getting or have received the COVID-19 vaccine. |
|  | Subjective norm 2 | Most of the people who are important to me (my family, relatives and/or friends) think I should get the COVID-19 vaccine. |
|  | Access | If I choose to get the COVID-19 vaccine, I believe it will be easy to get it. |
|  | Intention | I plan to get the COVID-19 vaccine. |
| COVID-19 testing and results, symptoms, and health behaviours | | |
|  | COVID symptoms | Since, January 2020, have you had any of the following symptoms? Check all that apply: headache, fever, stuffy nose/congestion, loss of smell or taste, new or worsening cough, difficulty breathing/shortness of breath, confusion, vomiting, chills, weakness, muscle pain, fatigue, nausea, diarrhea |
|  | Date of first symptoms | When did your first symptom start? (date) |
|  | Action following symptoms | Have you done any of the following for these symptoms? (Please check all that apply). Called family doctor/ GP, visited family doctor’s /GP office, visited community/public health clinic, been admitted to hospital etc. |
|  | Actions before symptoms | Before these symptoms, had you been in close contact with anyone who either: (A) had any of those symptoms [fever, new or worsening cough, headache, chills, weakness, muscle pain, stuffy nose/congestion, sore throat, difficulty breathing/shortness of breath, nausea, diarrhea, fatigue, loss of smell or taste, confusion, vomiting]; OR (B) was diagnosed positive for COVID-19 within 14 days before you felt sick? |
|  | Isolation before symptoms | Did you isolate, or stay away from your workplace or educational facility? |
|  | COVID-19 test | Have you been tested for COVID-19? |
|  | Test results | Did you test positive for COVID-19? |
|  | Household symptoms | Has anyone in your household either: (A) had any of the following symptoms: fever, new or worsening cough, headache, chills, weakness, muscle pain, stuffy nose/congestion, sore throat, difficulty breathing/shortness of breath, nausea, diarrhea, fatigue, loss of smell or taste, confusion, vomiting; OR (B) tested positive for COVID-19 since January 2020? |
|  | First symptoms date | When did their first symptom start? If you don’t remember, please make your best guess. |
|  | Household isolation | Has anyone in your household been told to quarantine, isolate, or limit time at their school or workplace since January 2020 because: they were sick or exposed to someone with COVID-19? |
|  | Adherence | Did they follow the advice and isolate, quarantine, or stay away from their workplace or educational facility? |
| Activities and behaviour in and outside of the home | | |
|  | Movement out of home | How many times did you leave your home (or property, apartment) yesterday? |
|  | Place of movement | Where did you go when you left your home? (Check all that apply) - Another person's home, a workplace, a hospital, doctor's office etc. |
|  | Distance | What is the farthest distance that you went from your home yesterday? |
|  | Means of transport | How did you travel when you left your home? (Check all that apply) - Selected Choice - I only walked (I did not use other transportation) |
|  | Face mask use | Did you use a face mask yesterday? |
|  | Face mask use location | Where did you use your face mask yesterday? (Check all that apply) - Selected Choice - Everywhere outside my house |
|  | Mask use duration | Take your best guess for the total amount of time you wore a mask yesterday (hours and minutes)? |
|  | Presence at home | In the last 3 hours, have you been in your home? |
|  | Handwashing | In the last 3 hours, have many times did you wash your hands with soap? |
|  | Hand sanitizer | In the last 3 hours, how many times did you use hand sanitizer? |
|  | Transport type | Yesterday, which type of public transportation did you use? (Please check all that apply) - Selected Choice - Airplane, bus, taxi etc |
|  | Transport duration | Yesterday, for about how long were you on public transportation? |
|  | PPE use during transportation | Yesterday, did you wear any of the following while on public transportation? (Please check all that apply) - Selected Choice - A face mask or other covering over your nose and mouth (e.g., face shield, bandana), gloves, etc. |
|  | Travel outside Canada | Have you travelled outside Canada at all since Jan 2020? And if so, to where? - Selected Choice |
|  | Number of contacts | Now we would like to ask you some questions about people you had in-person, face-to-face contact with yesterday. By in-person, face-to-face contact, we mean EITHER: A. An in-person two-way conversation with three or more words OR B. Physical skin-to-skin contact (for example, a handshake, hug, kiss, or contact sports). This includes family members, friends, co-workers, people you spoke to in shops, bus drivers, strangers, etc... and people of ALL ages. Please do not count people you contacted only with things like telephone, text, or online. How many people did you have in-person contact with between 5 am yesterday and 5 am today? |
|  | Contact identifier #1 to #10 | Please add a non-identifying "nickname" for each of the people you had face-to-face or physical contact with (e.g., DG, checkout person, bus driver, child #2). This "nickname" will help you to answer questions about this contact. - 1st person label |
|  | Characteristics of contact #1 (gender, age, relationship to respondent) | For the people you "nicknamed" and had in-person contact with between 5am yesterday and 5am today... - I believe this person identifies as… [indicate gender, age, relationship to you, location of contact, |
|  | Characteristics of contact #2 | Distance during contact, duration of contact, contact prior to COVID-19, PPE use during contact, distance during contact) |
|  | Location of contact of 10+ contacts | You said you had more than 10 in-person contacts. Where did majority of these contacts take place? |
|  | Occupational setting of 10+ contact | You said you had more than 10 in-person contacts. Which of these best describes your work/occupation or the other person's workplace where these contacts took place? |
|  | Age-group of contacts of 10+ contacts | You said you had more than 10 in-person contacts. What was the age-group for most of these contacts you interacted with? |
|  | Duration of 10+ contact | You said you had more than 10 in-person contacts. For most of these contacts, about how long did each contact last? |
| Internet and social media use and other information | | |
|  | Internet use | About how often do you use the internet? |
|  | Social media use | Thinking about the social media sites that you use; about how often do you visit or use each of the following? - Facebook, Instagram, Twitter, Snapchat, YouTube |
|  | Survey start date, survey end date, IP address, survey duration, response ID, recorded date, respondent's first and last name, location latitude, location longitude, follow up consent, draw consent | Survey start date, survey end date, IP address, survey duration, response ID, recorded date, respondent's first and last name, location latitude, location longitude, follow up consent, draw consent |
| **Derived variables** | | |
|  | Health Authority | The health authority of respondent. This was derived using respondents postal code or location data. |
|  | Quebec material index | The material deprivation involves deprivation of the goods and conveniences that are part of modern life, such as adequate housing, possession of a car, access to high-speed internet, or a neighbourhood with recreational areas. This deprivation marks the consequences of lack of material resources associated with low education, insecure job situation and insufficient income (1,2). |
|  | Quebec social index | Social deprivation refers to a fragile social network, starting with the family and encompassing the community. It is characterized by individuals living alone, being a lone parent and being separated, divorced, or widowed (1,2). |
|  | Ethnocultural composition | Ethno-cultural composition refers to the community make-up of immigrant populations, and at the British Columbia-level takes into consideration factors such as the proportion of population who self-identify as visible minority, the proportion of population that is foreign-born, the proportion of population with no knowledge of either official language (linguistic isolation), and the proportion of population who are recent immigrants (arrived in five years prior to Census). (1) |
|  | Situational vulnerability | Situational vulnerability speaks to variations in socio-demographic conditions in the areas of housing and education, while taking into account other demographic characteristics. The indicators in this dimension at the British Columbia-level measure concepts such as the proportion of population that identifies as Aboriginal, the proportion of population aged 25-64 without a high school diploma, the proportion of dwellings needing major repairs, the proportion of population that is low-income, and the proportion of single parent families (1). |
|  | Economic dependency | Economic dependency relates to reliance on the workforce, or a dependence on sources of income other than employment income. Indicators included in this dimension at the British Columbia-level measure concepts such as the proportion of population participating in labour force (aged 15 and older), the proportion of population aged 65 and older, the ratio of employment to population, and the dependency ratio (population aged 0-14 and aged 65 and older divided by population aged 15-64)(1). |
|  | Residential instability | Residential instability speaks to the tendency of neighbourhood inhabitants to fluctuate over time, taking into consideration both housing and familial characteristics. The indicators in this dimension at the British Columbia-level measure concepts such as the proportion of dwellings that are apartment buildings, the proportion of persons living alone, the proportion of dwellings that are owned, and the proportion of the population who moved within the past five years (1). |

| **S2 Table.** Comparison of baseline eligible and ineligible participants, frequencies and proportions | | | | | | | |  |
| --- | --- | --- | --- | --- | --- | --- | --- | --- |
|  |  | Eligible (n=41,375) | | |  | Ineligible (n=15, 702) **^‡^** | | |
|  |  | Frequency | Percent (incl. missing) | Percent (excl. missing) |  | Frequency | Percent (incl. missing) | Percent (excl. missing) |
| Sex |  |  |  |  |  |  |  |  |
|  | Male | 6,823 | 16.5 | 16.5 |  | 1,697 | 10.8 | 18.6 |
|  | Female | 34,552 | 83.5 | 83.5 |  | 7,442 | 47.4 | 81.4 |
|  | Missing | n/a | n/a | n/a |  | 6,563 | 41.8 | n/a |
| Age |  |  |  |  |  |  |  |  |
|  | 18-34 | 4,978 | 12.0 | 12.0 |  | 1,726 | 11.0 | 17.7 |
|  | 35-54 | 12,110 | 29.3 | 29.3 |  | 3,039 | 19.4 | 31.2 |
|  | 55+ | 24,287 | 58.7 | 58.7 |  | 4,981 | 31.7 | 51.1 |
|  | Missing | n/a | n/a | n/a |  | 5,956 | 37.9 | n/a |
| Race/ethnicity |  |  |  |  |  |  |  |  |
|  | Indigenous | 1,757 | 4.3 | 4.4 |  | 666 | 4.2 | 7.2 |
|  | Chinese | 882 | 2.1 | 2.2 |  | 238 | 1.5 | 2.6 |
|  | White | 35,026 | 84.7 | 87.5 |  | 7,439 | 47.4 | 79.9 |
|  | South Asian | 606 | 1.5 | 1.5 |  | 315 | 2.0 | 3.4 |
|  | Other | 1,766 | 4.3 | 4.4 |  | 649 | 4.1 | 7.0 |
|  | Missing/Unknown | 1,338 | 3.2 | n/a |  | 6,395 | 40.7 | n/a |
| Health region |  |  |  |  |  |  |  |  |
|  | Fraser Health | 8,451 | 20.4 | 26.1 |  | 1,802 | 11.5 | 31.0 |
|  | Interior Health | 6,143 | 14.8 | 19.0 |  | 1,061 | 6.8 | 18.3 |
|  | Northern Island | 1,825 | 4.4 | 5.6 |  | 312 | 2.0 | 5.4 |
|  | Vancouver Coastal | 7,315 | 17.7 | 22.6 |  | 1,329 | 8.5 | 22.9 |
|  | Vancouver Island | 8,640 | 20.9 | 26.7 |  | 1,300 | 8.3 | 22.4 |
|  | Missing | 9,001 | 21.8 | n/a |  | 9,898 | 63.0 | n/a |
| Education |  |  |  |  |  |  |  |  |
|  | Below high school | 807 | 2.0 | 2.5 |  | 41 | 0.3 | 7.1 |
|  | Below bachelor | 16,928 | 40.9 | 51.7 |  | 245 | 1.6 | 42.5 |
|  | University degree | 15,029 | 36.3 | 45.9 |  | 290 | 1.8 | 50.3 |
|  | Missing/Unknown | 8,611 | 20.8 | n/a |  | 15,126 | 96.3 | n/a |
| Employment status |  |  |  |  |  |  |  |  |
|  | Employed full-time (30 hours or more/week) | 10,654 | 25.7 | 32.0 |  | 210 | 1.2 | 31.6 |
|  | Employed part-time | 2,993 | 7.2 | 9.0 |  | 75 | 0.4 | 11.3 |
|  | Self-employed | 2,704 | 6.5 | 8.1 |  | 64 | 0.4 | 9.6 |
|  | Unemployed but looking for a job | 952 | 2.3 | 2.9 |  | 24 | 0.1 | 3.6 |
|  | Unemployed and not looking for a job | 406 | 1.0 | 1.2 |  | 10 | 0.1 | 1.5 |
|  | Full-time parent, homemaker | 879 | 2.1 | 2.6 |  | 10 | 0.1 | 1.5 |
|  | Retired | 12,757 | 30.8 | 38.3 |  | 87 | 0.5 | 13.1 |
|  | Student/Pupil | 566 | 1.4 | 1.7 |  | 67 | 0.4 | 10.1 |
|  | Long-term sick or disabled | 968 | 2.3 | 2.9 |  | 31 | 0.2 | 4.7 |
|  | Prefer not to answer | 424 | 1.0 | 1.3 |  | 87 | 0.5 | 13.1 |
|  | Missing/Unknown | 8,072 | 19.5 | n/a |  | 15,037 | 84.8 | n/a |
| Quebec Material Deprivation Index |  |  |  |  |  |  |  |  |
|  | 1 (Privileged) | 6,407 | 15.5 | 22.3 |  | 690 | 4.4 | 13.8 |
|  | 2 | 6,475 | 15.6 | 22.5 |  | 1,041 | 6.6 | 20.9 |
|  | 3 | 6,972 | 16.9 | 24.2 |  | 1,538 | 9.8 | 30.8 |
|  | 4 | 4,822 | 11.7 | 16.8 |  | 751 | 4.8 | 15.1 |
|  | 5 (Deprived) | 4,085 | 9.9 | 14.2 |  | 969 | 6.2 | 19.4 |
|  | Missing | 12,614 | 30.5 | n/a |  | 10,713 | 68.2 | n/a |
| Quebec Social Deprivation Index |  |  |  |  |  |  |  |  |
|  | 1 (Privileged) | 4,932 | 11.9 | 17.2 |  | 1,018 | 6.5 | 20.4 |
|  | 2 | 4,756 | 11.5 | 16.5 |  | 696 | 4.4 | 14.0 |
|  | 3 | 6,311 | 15.3 | 21.9 |  | 1,275 | 8.1 | 25.6 |
|  | 4 | 5,932 | 14.3 | 20.6 |  | 897 | 5.7 | 18.0 |
|  | 5 (Deprived) | 6,830 | 16.5 | 23.8 |  | 1,103 | 7.0 | 22.1 |
|  | Missing | 12,614 | 30.5 | n/a |  | 10,713 | 68.2 | n/a |
| Follow up consent |  |  |  |  |  |  |  |  |
|  | Yes | 20,633 | 49.9 | 63.8 |  | 245 | 1.6 | 39.6 |
|  | No | 11,689 | 28.3 | 36.2 |  | 373 | 2.4 | 60.4 |
|  | Missing | 9,053 | 21.9 | n/a |  | 15,084 | 96.1 | n/a |
| Data linkage consent |  |  |  |  |  |  |  |  |
|  | Yes | 7,290 | 17.6 | 27.3 |  | 95 | 0.6 | 17.3 |
|  | No | 19,467 | 47.1 | 72.8 |  | 454 | 2.9 | 82.7 |
|  | Missing | 14,618 | 35.3 | n/a |  | 15,153 | 96.5 | n/a |
| **^‡^**Does not include 4,106 duplicates | | | | | | | | |

**S3 Table.** Participant profile of BC-Mix follow up surveys: frequencies and proportions (%)

|  | Follow up#1 (n=10,993) | Follow up #2 (n=8,164) | Follow up #3 (n=6,375) | Follow up #4 (n=4,981) | Follow up #5 (n=3,891) | Follow up #6 (n=3,184) | Follow up #7 (n=2,417) | Follow up #8 (n=1,760) |
| --- | --- | --- | --- | --- | --- | --- | --- | --- |
| **Sex** | | | | | | | | |
| Male | 1590 (14.5) | 1115 (13.7) | 843 (13.2) | 646 (13.0) | 495 (12.7) | 404 (12.17) | 312 (12.9) | 217 (12.3) |
| Female | 9403 (85.5) | 7049 (86.3) | 5532 (86.8) | 4335 (87.0) | 3396 (87.3) | 2780 (87.3) | 2105 (87.1) | 1543 (87.7) |
| **Age** | | | | | | | | |
| 18-34 | 1128 (10.3) | 731 (9.0) | 497 (7.8) | 363 (7.3) | 257 (6.6) | 199 (6.3) | 152 (6.3) | 109 (6.2) |
| 35-54 | 3013 (27.4) | 2105 (25.8) | 1533 (24.1) | 1127 (22.6) | 846 (21.7) | 662 (20.8) | 495 (20.5) | 343 (19.5) |
| 55+ | 6852 (62.3) | 5328 (65.3) | 4345 (68.2) | 3491 (70.1) | 2788 (71.7) | 2323 (73.0) | 1770 (73.2) | 1308 (74.3) |
| **Race/ethnicity** | | | | | | | | |
| Indigenous | 342 (3.1) | 229 (2.8) | 162 (2.5) | 130 (2.6) | 91 (2.3) | 71 (2.2) | 54 (2.2) | 38 (2.2) |
| Chinese | 199 (1.8) | 124 (1.5) | 98 (1.5) | 66 (1.3) | 48 (1.2) | 39 (1.2) | 25 (1.0) | 16 (0.9) |
| White | 9870 (89.8) | 7415 (90.8) | 5833 (91.5) | 4586 (92.1) | 3602 (92.6) | 2959 (92.9) | 2254 (93.3) | 1642 (93.3) |
| South Asian | 79 (0.7) | 39 (0.5) | 30 (0.5) | 20 (0.4) | 15 (0.4) | 14 (0.4) | 12 (0.5) | 12 (0.7) |
| Other | 316 (2.9) | 229 (2.8) | 154 (2.4) | 108 (2.2) | 86 (2.2) | 63 (2.0) | 45 (1.9) | 35 (2.0) |
| Missing/Unknown | 187(1.7) | 128 (1.6) | 98 (1.5) | 71 (1.4) | 49 (1.3) | 38 (1.2) | 27 (1.1) | 17 (1.0) |
| **Health region** | | | | | | | | |
| Fraser Health | 2748 (25.0) | 2039 (25.0) | 1590 (24.9) | 1243 (25.0) | 964 (24.8) | 792 (24.9) | 613 (25.4) | 462 (26.3) |
| Interior Health | 1926 (17.5) | 1435 (17.6) | 1156 (18.1) | 907 (18.2) | 703 (18.1) | 579 (18.2) | 432 (17.9) | 317 (18.0) |
| Northern Island | 506 (4.6) | 374 (4.6) | 280 (4.4) | 212 (4.3) | 162 (4.2) | 116 (3.4) | 88 (3.6) | 63 (3.6) |
| Vancouver Coastal | 2706 (24.6) | 1992 (24.4) | 1540 (24.2) | 1178 (23.7) | 932 (24.0) | 758 (23.8) | 577 (23.9) | 403 (22.9) |
| Vancouver Island | 3059 (27.8) | 2303 (28.2) | 1794 (28.1) | 1430 (28.7) | 1122 (28.8) | 934 (29.3) | 703 (29.1) | 513 (29.2) |
| Missing/Unknown | 48 (0.4) | 21 (0.3) | 15 (0.2) | 11 (0.2) | 8 (0.2) | 5 (0.2) | 4 (0.2) | 2 (0.1) |
| **Education** | | | | | | | | |
| Below high school | 173 (1.5) | 123 (1.5) | 89 (1.4) | 68 (1.4) | 51 (1.3) | 37 (1.2) | 26 (1.1) | 16 (0.9) |
| Below bachelor | 5236 (47.6) | 3835 (47.0) | 2979 (46.7) | 2303 (46.2) | 1771 (45.5) | 1453 (45.6) | 1108 (45.8) | 802 (45.6) |
| University degree | 5529 (50.3) | 4169 (51.1) | 3283 (51.5) | 2594 (52.1) | 2057 (52.9) | 1683 (52.9) | 1278 (52.9) | 939 (53.4) |
| Missing/Unknown | 55 (0.5) | 37 (0.5) | 24 (0.4) | 16 (0.3) | 12 (0.3) | 11 (0.4) | 5 (0.2) | 31 (0.2) |
| **Quebec Material Deprivation Index** | | | | | | | | |
| 1 (Privileged) | 1547 (14.1) | 1072 (13.75) | 896 (14.1) | 720 (14.5) | 569 (14.6) | 435 (13.7) | 344 (14.2) | 253 (14.4) |
| 2 | 2078 (18.9) | 1547 (19.0) | 1198 (18.8) | 1008 (20.2) | 806 (20.7) | 665 (20.9) | 500 (20.7) | 366 (20.8) |
| 3 | 2994 (27.3) | 2184 (26.8) | 1674 (26.3) | 1273 (25.6) | 973 (25.0) | 835 (26.2) | 642 (26.6) | 450 (25.6) |
| 4 | 1277 (11.6) | 942 (11.64) | 748 (11.7) | 580 (11.6) | 475 (12.2) | 379 (11.9) | 285 (11.8) | 215 (12.2) |
| 5 (Deprived) | 1668 (15.2) | 1290 (15.8) | 1033 (16.2) | 728 (15.7) | 609 (15.7) | 504 (15.8) | 375 (15.5) | 278 (15.8) |
| Missing | 1429 (13.0) | 1069 (13.1) | 826 (13.0) | 616 (12.4) | 459 (11.8) | 366 (11.5) | 271 (11.2) | 198 (11.3) |
| **Quebec Social Deprivation Index** | | | | | | | | |
| 1 (Privileged) | 2188 (19.9) | 1641 (20.1) | 1286 (20.2) | 1063 (21.3) | 830 (20.3) | 658 (20.7) | 509 (21.1) | 368 (20.9) |
| 2 | 1441 (13.1) | 1051 (12.9) | 787 (12.4) | 603 (12.1) | 480 (12.3) | 411 (12.9) | 314 (13.0) | 225 (12.8) |
| 3 | 2478 (22.5) | 1831 (22.4) | 1480 (23.2) | 1148 (23.1) | 922 (23.7) | 769 (24.2) | 574 (23.8) | 419 (23.8) |
| 4 | 1601 (14.6) | 1236 (15.1) | 945 (14.8) | 743 (14.9) | 578 (14.9) | 488 (15.3) | 370 (15.3) | 290 (16.5) |
| 5 (Deprived) | 1856 (16.9) | 1336 (16.4) | 1051 (16.5) | 808 (16.2) | 622 (16.0) | 392 (15.5) | 379 (15.7) | 260 (14.8) |
| Missing/Unknown | 1429 (13) | 1069 (13.1) | 826 (13.0) | 616 (12.4) | 459 (11.8) | 492 (11.5) | 271 (11.2) | 198 (11.3) |
| **Follow up consent** | | | | | | | | |
| Yes | 10357 (94.2) | 7793 (95.5) | 6182 (97.0) | 4857 (97.5) | 3789 (97.4) | 3106 (97.6) | 2380 (98.5) | 1714 (97.4) |
| No | 262 (2.4) | 142 (1.7) | 83 (1.3) | 49 (1.0) | 47 (1.2) | 31 (1.0) | 17 (0.7) | 19 (1.1) |
| Missing | 374 (3.4) | 229 (2.8) | 110 (1.7) | 75 (1.5) | 55 (1.4) | 47 (1.5) | 20 (0.8) | 27 (1.5) |
